# Kidney disease screening among adults with HIV in Uganda: a missed priority for a high-risk population

**DOI:** 10.1101/2025.08.08.25333274

**Authors:** Grace Kansiime, Joseph Baruch Baluku, Edwin Nuwagira, Michael Kanyesigye, Paul Stephen Obwoya, Rose Muhindo, Winnie R. Muyindike, Pliers Denis Tusingwire, Henry Mugerwa, Matthew Odera, Monday Busuulwa, Matthew Ssemakadde, Esther C. Atukunda, Francis Bajunirwe, Robert Kalyesubula, Mark J. Siedner

**Author notes:** **Corresponding Author and Address:** Grace Kansiime, Department of Medicine, Mbarara University of Science and Technology, PO Box 1410, Mbarara, Uganda.

## Abstract

Chronic kidney disease affects 850 million people worldwide, with Sub-Saharan Africa bearing a significant burden. People living with HIV (PWH) are at increased risk due to nephrotoxicity of antiretroviral therapy (ART), in part due to widespread use of tenofovir disoproxil fumarate. In response, Uganda recommends routine kidney disease screening at ART initiation. However, the extent of adherence to these guidelines remains poorly understood.

We extracted clinical data for adults initiating ART between 2017 and 2024 at three large-volume HIV clinics in Uganda. To determine if kidney disease screening rates had increased appropriately over time, we divided the observation period into three eras as per national guidelines: (1) Test and Treat (2017-2019), that recommended screening only PWH and diabetes or hypertension; (2) DTG rollout/COVID-19 (2020-2022); and (3) creatinine-for-all (2023-2024), recommending screening everyone initiating ART. Logistic regression models were fit to identify correlates of renal screening.

Of the 17,485 participants, only 22.4% (3,909/17,485) were screened for kidney disease. Screening was more common at the urban site (54.2%) compared to rural sites (10.0%). At rural sites, screening declined over time and individuals were 83% less likely to be screened in the creatinine-for-all era compared to the baseline era (aOR 0.17, 95% CI: 0.13–0.22) while it increased at urban site (aOR 9.27, 95% CI: 7.37–11.66). Male sex (aOR 1.37, 95% CI: 1.20– 1.57), older age (>45 years), hypertension, and non–TDF-based ART regimens were associated with higher screening odds at rural sites. Diabetes, opportunistic infections, and TDF use were not significantly associated with screening likelihood at any site.

Kidney disease screening at ART initiation remains poor in Uganda, particularly in rural clinics, highlighting critical challenges in translating national guidelines into practice. Future research should focus on understanding multilevel barriers to screening and evaluating strategies to improve guideline uptake.

## Background

Chronic kidney disease (CKD) is a growing challenge globally, affecting 850 million people worldwide and contributing significantly to morbidity and mortality.(1-4) Sub-Saharan Africa (SSA) bears a disproportionately high burden of CKD, with prevalence estimates exceeding 15%.(5-7) The increased rates of CKD in SSA are attributed to rising prevalence of diabetes and hypertension, regionally common genetic predispositions such as sickle cell trait and APOL 1, and endemic infections such as tuberculosis, malaria, schistosomiasis, hepatitis B and C, and HIV.(7-12)

SSA bears the highest global burden of HIV. Over half of the people living with HIV (PWH) worldwide reside in eastern and southern Africa.(13, 14) PWH are at increased risk for CKD due to direct pathogenicity of the HIV and nephrotoxicity of antiretroviral therapy (ART) agents, notably tenofovir disoproxil fumarate (TDF), the main backbone of ART in SSA.(15-18) Late diagnosis and referral for nephrology care among PWH may worsen the prognosis of CKD, leading to complications of cardiovascular disease, bone disorders, dyslipidemia, cognitive decline, and decreased quality of life.(19-23)

In response, many countries, including Uganda, recommend routine kidney disease screening before starting ART as well as regular monitoring while on ART.(24) However, whether such guidelines are followed in people initiating ART in the region is not well studied. We hypothesized that kidney disease screening remains low for PWH in Uganda, despite the advancement of guidelines to do so. To test this hypothesis, we evaluated routine kidney disease screening among PWH in Uganda over the past eight years (2017-2024) in three large-volume HIV clinics in Uganda.

## Methods

### Study Setting and Participants

We conducted data analysis for a retrospective cohort of PWH on ART at three large volume HIV clinics in Uganda: Mbarara Regional Referral Immune Suppression (ISS) clinic, Masaka Regional Referral Hospital (RRH) HIV clinic as rural clinics, and Joint Clinical Research Centre (JCRC) HIV clinic an urban clinic, with combined patient census of 50,000. We included all adults aged 18 years and above, initiated on ART between January 2017 and December 2024.

### Data collection methods

All three clinics have electronic medical records systems that record patients’ clinical encounters, medications prescribed, and results of laboratory tests. We extracted demographic and clinical data for all adult patients who started ART during the observation period. Extracted indicators included; ART initiation date, creatinine test dates and results, age at ART initiation, sex, education level, marital status, occupation, weight, height, alcohol use and smoking history, co-morbidity history (hypertension, diabetes, tuberculosis, cryptococcal meningitis, and Kaposi sarcoma), CD4 count at ART initiation, and baseline ART regimen.

### Statistical methods

Data were analyzed using Stata software, version SE 17.1. We first summarized continuous variables using medians and interquartile ranges (IQR), and categorical variables using percentages. We subsequently examined changes in kidney disease screening over time in Uganda. Our primary outcome of interest was completion of kidney disease screening, defined as having a serum creatinine test done within three months of ART initiation. Our primary exposure of interest was the ART initiation period, which we defined a priori as three era:. 1) the “test and treat” era, where all PWH were eligible for ART regardless of CD4 count and creatinine screening was recommended for those with comorbid diabetes or hypertension; 2) the “DTG and COVID-19”, era where all individuals were started on TLD, but health systems were constrained due to the COVID-19 pandemic; and 3) the “creatinine screening for all” era when creatinine screening for all individuals starting ART was first implemented. (Table 1)

**Table 1.** Study time periods of interest.

| Years | 2017-2019 | 2020-2022 | 2023-2024 |
| --- | --- | --- | --- |
| Study Period | Test and Treat era | DTG and Covid-19 era | Creatinine for all era |
| **Uganda ART Guidelines and other era considerations** | All PWH eligible for therapy. First-line regimen: efavirenz, tenofovir, and lamivudine | All PWH eligible for therapy. First-line regimen: dolutegravir, tenofovir, and lamivudine. Some care interruptions due to COVID-19 lockdowns | All PWH eligible for therapy. First-line regimen: dolutegravir, tenofovir, and lamivudine. |
| **Uganda kidney disease screening guidelines for PWH** | Serum creatinine for PWH with comorbid diabetes or hypertension.(25) | Serum creatinine for PWH with comorbid diabetes or hypertension.(26) | Serum creatinine screening for all PWH at time of ART initiation.(24) |

*ART, Antiretroviral therapy; DTG, dolutegravir; PWH, people with HIV*

Crude rates of kidney disease screening were estimated by clinic type and period. Multivariable logistic regression models were utilized to estimate the association between selected covariates and kidney disease screening at ART initiation. Notably, a significant qualitative interaction was observed between creatinine screening changes over time and the urban (JCRC) versus rural (Mbarara and Masaka) sites. Consequently, these were treated as distinct populations.

### Human subjects and Ethical considerations

The study was reviewed and approved by the Mbarara University Review and Ethics Committee (MUST ID: 2024-1667) and received a waiver of informed consent. Additionally, we obtained approval from the Uganda National Council of Science and Technology (HS5305ES), and administrative clearances from all the study sites.

## Results

### Study population

Between January and May 2025, we extracted data for 17,485 adults who were initiated on ART from January 2017 to December 2024 at Mbarara RRH, Masaka RRH, and JCRC HIV clinics in Uganda. The baseline and clinical characteristics of participants were largely similar across the three ART initiation periods (Table 2). Most participants (n=11,068, 63%) were female, and the majority (n=14,907, 85.2%) were younger than 45 years of age, The median CD4 count was 378 (interquartile range [IQR] 172-631) cells/uL, and less than 10% (n=1,319) had diabetes or hypertension. For those with available sociodemographic data, about half (n=6,349, 54.9%) had a primary level education or less. Most (n=5,532, 88.9%) reported being employed.

**Table 2.** Participant baseline characteristics

|  | Study period<br>Test & treat era<br>n=8,164 | DTG/COVID-19 era<br>n=5,764 | Creatinine for all era<br>n=3,557 | Total<br>N=17,485 |
| --- | --- | --- | --- | --- |
| **Sex** |  |  |  |  |
| Male | 2,921 (35.8%) | 2,118 (36.7%) | 1,378 (38.7%) | 6,417 (36.7%) |
| Female | 5,243 (64.2%) | 3,646 (63.3%) | 2,179 (61.3%) | 11,068 (63.3%) |
| **Age at ART start, median (IQR)** | 30 (25-38) | 32 (26-40) | 32 (25.3-40) | 31 (25-39) |
| **Age category (years)** |  |  |  |  |
| 18-24 | 2,040 (25.0%) | 1,186 (20.6%) | 766 (21.5%) | 3,992 (22.8%) |
| 25-44 | 5,103 (62.5%) | 3,594 (62.4%) | 2,218 (62.4%) | 10,915 (62.4%) |
| ≥ 45 | 1,021 (12.5%) | 984 (17.1%) | 573 (16.1%) | 2,578 (14.7%) |
| **Education n=11,572** |  |  |  |  |
| Primary and below | 3,567 (56.9%) | 1,955 (51.8%) | 827 (53.9%) | 6,349 (54.9%) |
| Secondary | 2,162 (34.5%) | 1,395 (37.0%) | 495 (32.2%) | 4,052 (35.0%) |
| Tertiary | 537 (8.6%) | 421 (11.2%) | 213 (13.9%) | 1,171 (10.1%) |
| **Study site** |  |  |  |  |
| ISS Mbarara | 3,009 (36.9%) | 1,792 (31.1%) | 1,001 (28.1%) | 5,802 (33.2%) |
| Masaka | 3,609 (44.2%) | 1,998 (34.7%) | 1,069 (30.1%) | 6,676 (38.2%) |
| JCRC | 1,546 (18.9%) | 1,974 (34.2%) | 1,487 (41.8%) | 5,007 (28.6%) |
| **Occupation, n=12,052** |  |  |  |  |
| Unemployed | 690 (11.1%) | 364 (9.8%) | 157 (7.4%) | 1,211 (10.0%) |
| Business | 1,897 (30.5%) | 1,169 (31.4%) | 736 (34.9%) | 3,802 (31.5%) |
| Farmer | 1,318 (21.2%) | 683 (18.4%) | 309 (14.6%) | 2,310 (19.2%) |
| Other | 2,317 (37.2%) | 1,504 (40.4%) | 908 (43.0%) | 4,729 (39.2%) |
| **Marital status n= 8,494** |  |  |  |  |
| Not married | 1,831 (47.9%) | 1,463 (50.6%) | 980 (55.0%) | 4,274 (50.3%) |
| Married | 1,991 (52.1%) | 1,428 (49.4%) | 801 (45.0%) | 4,220 (49.7%) |
| **Baseline CD4 (cell/μL)**<br>**median (IQR)** | 389 (181-666) | 378 (175-624) | 359 (148-597) | 378 (172-631) |
| **CD4 category n=10,166** |  |  |  |  |
| <200 | 1,050 (26.8%) | 1,011 (28.4%) | 812 (30.2%) | 2,873 (28.3%) |
| 200-500 | 1,334 (34.0%) | 1,271 (35.7%) | 958 (35.6%) | 3,563 (35.0%) |
| >500 | 1,535 (39.2%) | 1,277 (35.9%) | 918 (34.2%) | 3,730 (36.7%) |
| **Comorbidities** |  |  |  |  |
| Hypertension | 459 (5.6%) | 288 (5.0%) | 164 (4.6%) | 911 (5.2%) |
| Diabetes mellitus | 171 (2.1%) | 155 (2.7%) | 82 (2.3%) | 408 (2.3%) |
| Tuberculosis | 635 (7.8%) | 533 (9.2%) | 343 (9.6%) | 1,511 (8.6%) |
| Cryptococcal meningitis | 145 (1.8%) | 128 (2.2%) | 31 (0.9%) | 304 (1.7%) |
| Kaposi sarcoma | 92 (1.1%) | 48 (0.8%) | 10 (0.3%) | 150 (0.9%) |
| **Baseline Regimen n= 16,212** |  |  |  |  |
| TDF based | 7,423 (97.9%) | 5,180 (98.3%) | 3,317 (98.7%) | 15,920 (98.2%) |
| Non-TDF based | 158 (2.1%) | 91 (1.7%) | 43 (1.3%) | 292 (1.8%) |

ART, Antiretroviral therapy; CD4, Cluster of differentiation; DM, diabetes mellitus; DTG, dolutegravir; HTN, hypertension; ISS, Immune suppression syndrome, IQR, interquartile range, JCRC, Joint clinical research centre; PWH, people with HIV; TDF, tenofovir disoproxil fumarate; TLD, Tenofovir Lamivudine Dolutegravir; TLE, Tenofovir Lamivudine, Efavirenz; yrs, years

### Prevalence of kidney disease screening in Uganda

The proportion of the total population who completed kidney disease screening within three months of ART initiation during the observation period was 22.4% (3,909/17,485, 95%CI 21.7 – 23.0). At the urban site, 54.2% (95%CI 52.8-55.6) completed kidney disease screening compared to 9.6% (95%CI 9.1-10.1) at the rural sites (Fig 1).

**Fig 1:**
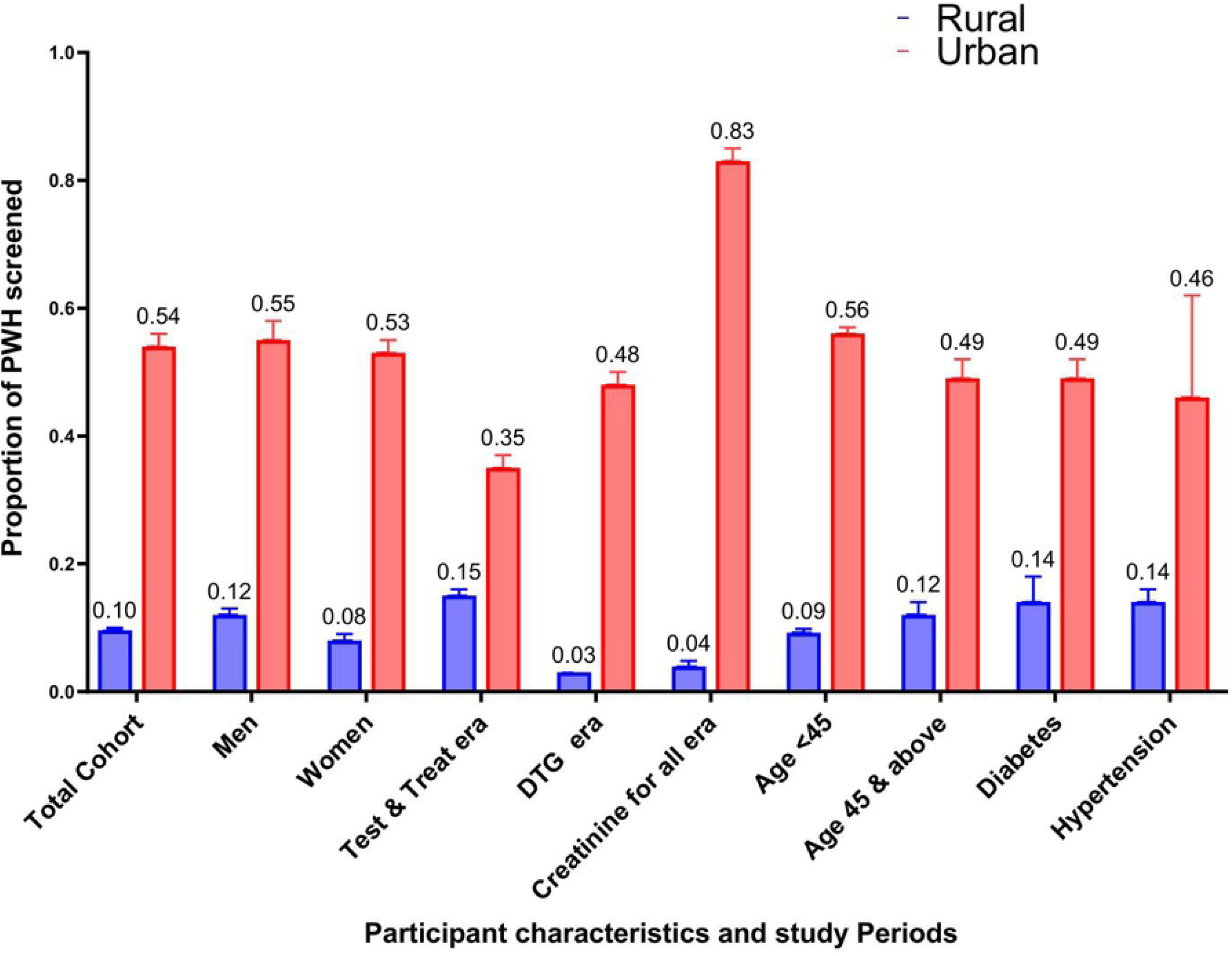
Proportion of PWH screened for kidney disease at ART initiation at rural and urban site

Similarly, the crude occurrence of kidney disease screening was consistently higher over the study observation at the urban site, while it declined or remained stagnant at the rural sites (Fig 2).

**Fig 2:**
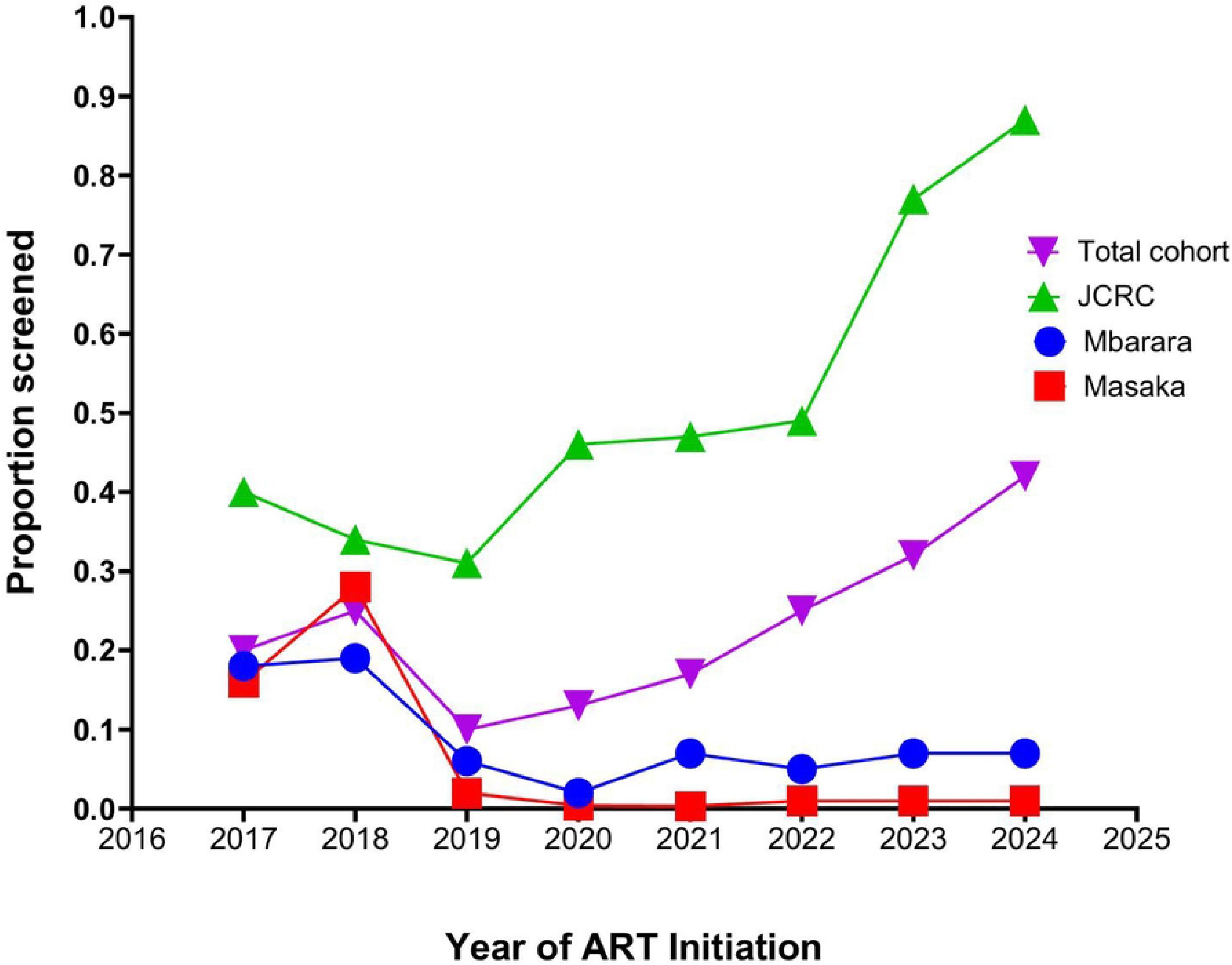
Kidney disease screening trends for PWH in Uganda by ART initiation year

### Factors associated with kidney disease screening in Uganda

At rural study sites, kidney disease screening rates declined over time. Compared to the initial test and treat period, participants initiating ART in the creatinine-for-all era had an 83% lower likelihood of kidney disease screening (aOR 0.17, 95% CI: 0.13 - 0.22, p <0.001, Table 3A). In contrast, the urban site showed a substantial increase in screening during the creatinine-for-all period, with participants over nine times more likely to be screened than in the test and treat era (aOR 9.27, 95% CI: 7.37-11.66, p <0.001, Table 3B).

**Table 3A.**
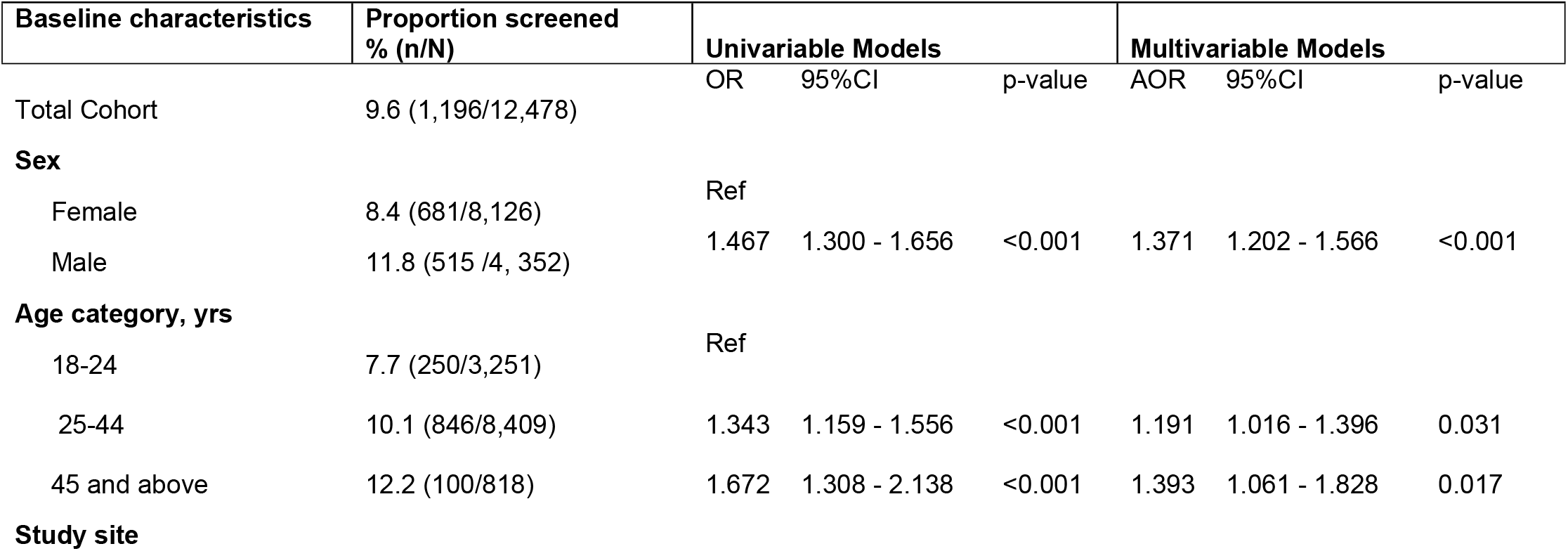
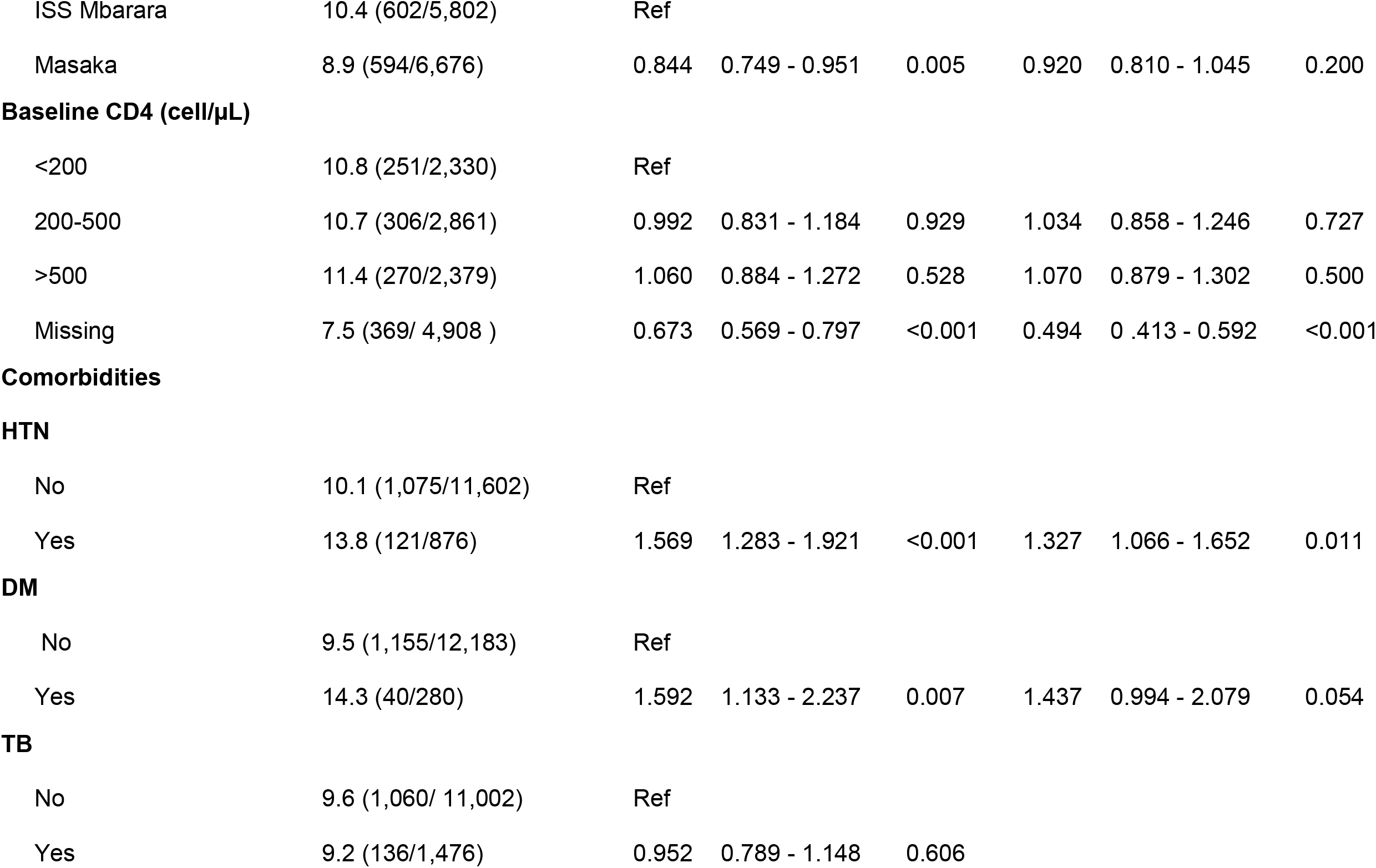
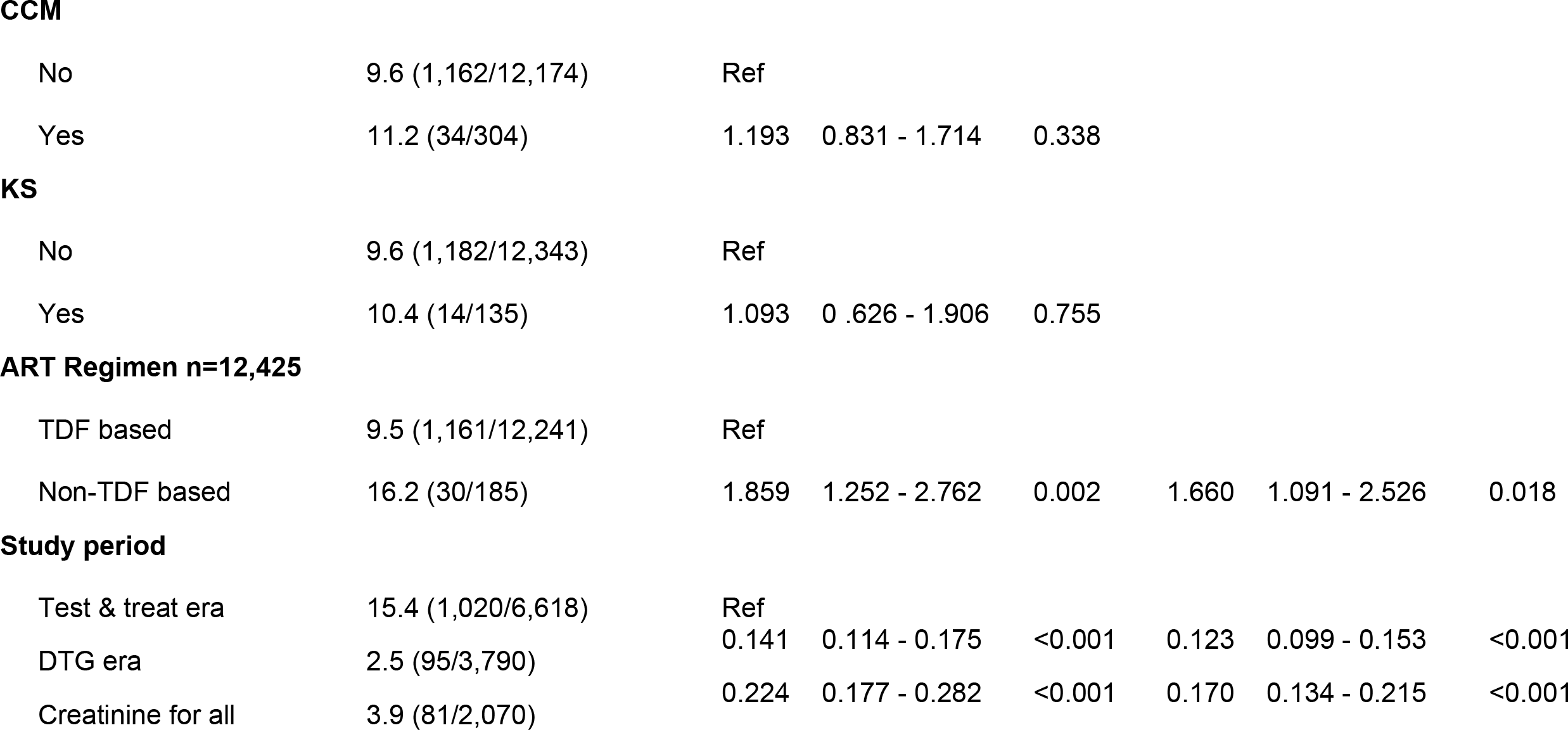
Correlates of kidney screening at rural Uganda clinics

**Table 3B.**
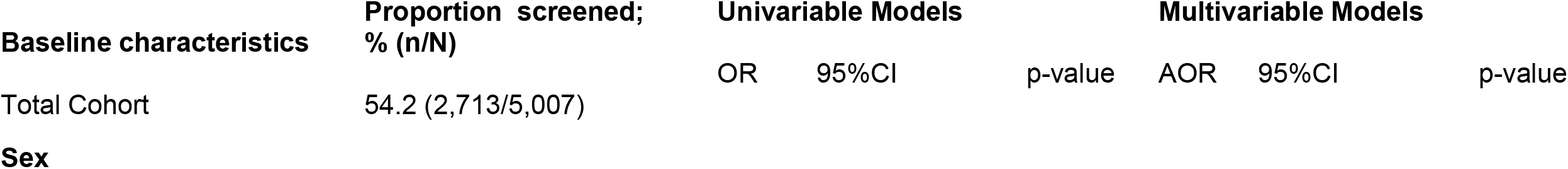
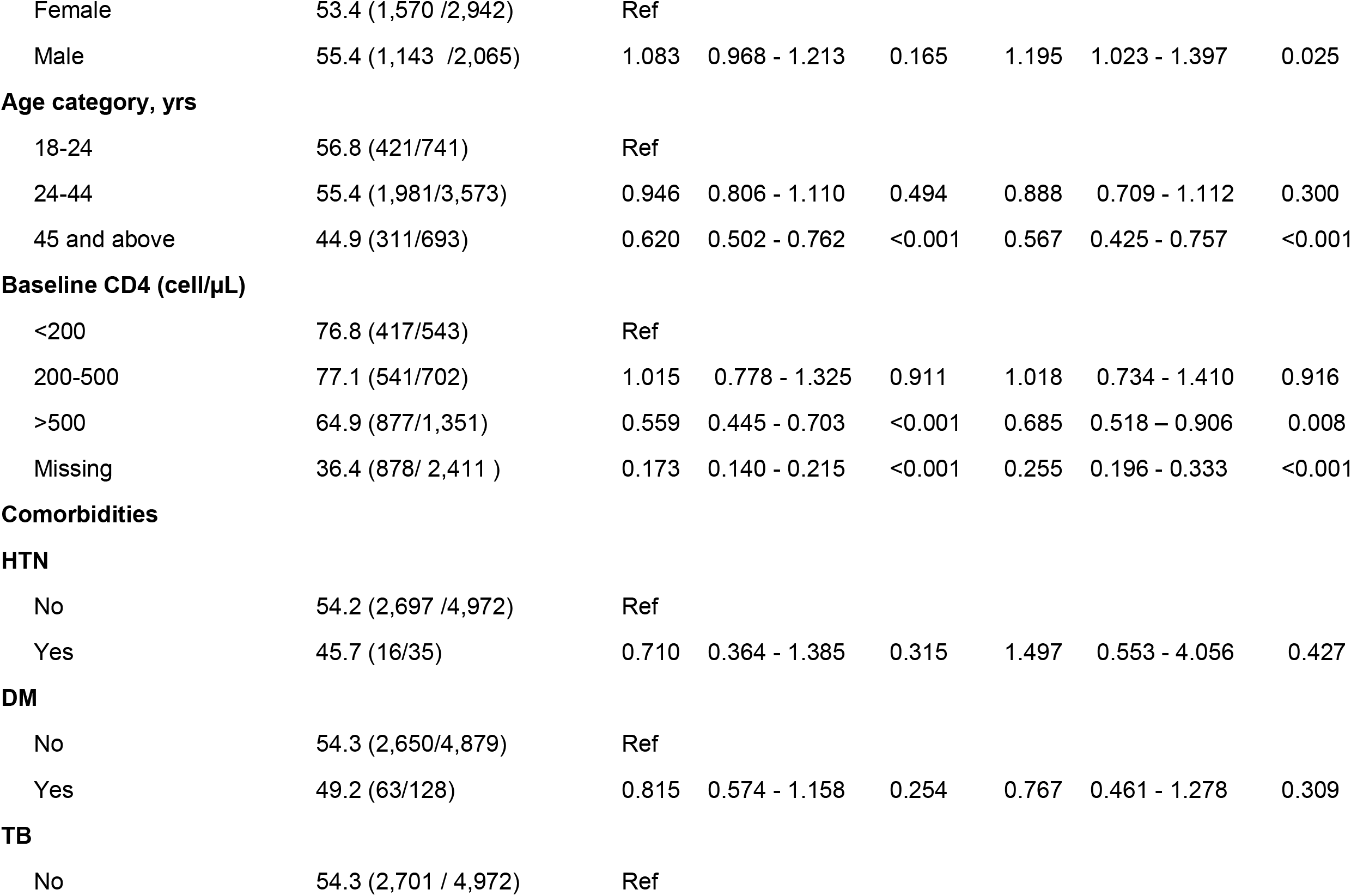
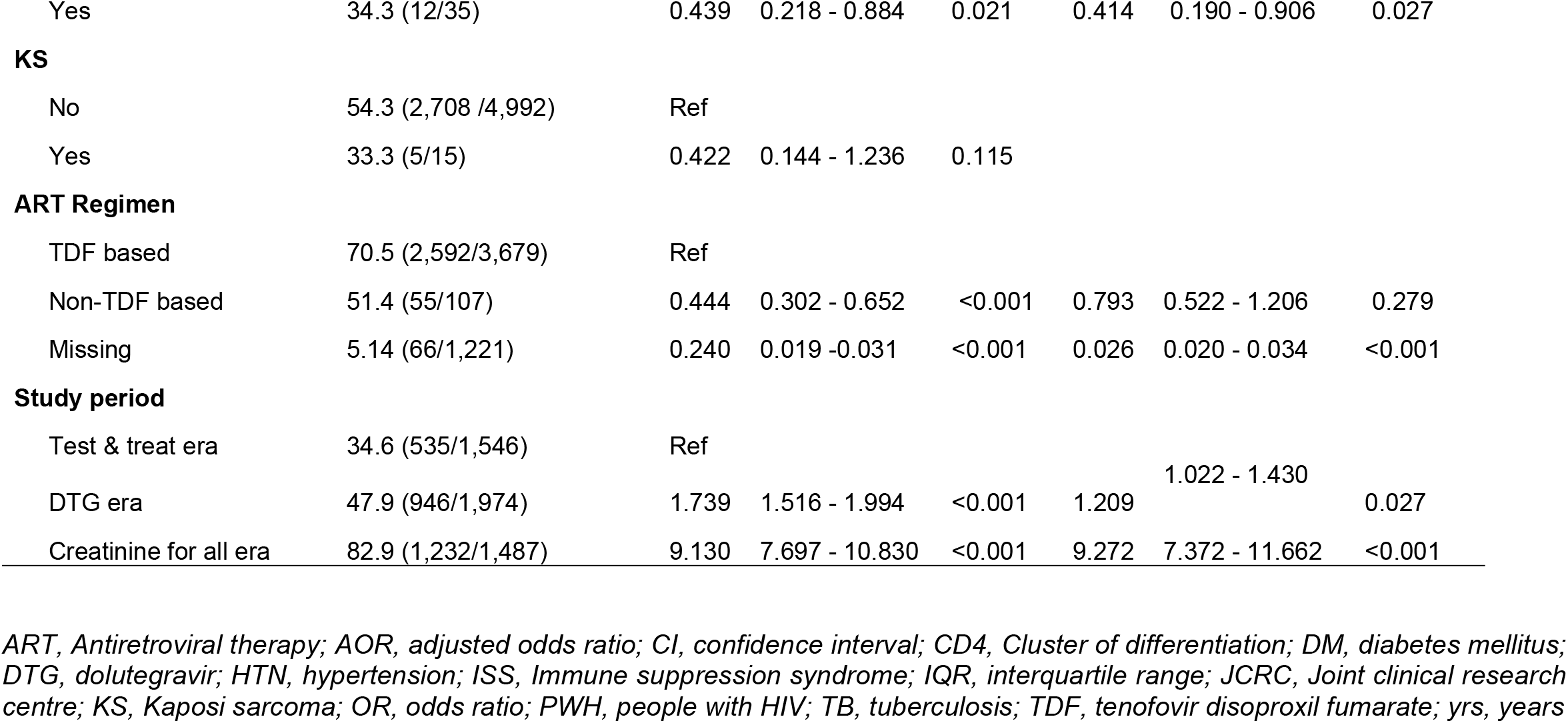
Correlates of kidney screening at a large urban Ugandan clinic

Several factors were associated with higher odds of kidney disease screening at ART initiation in rural sites (Table 3A). For example, male participants were about 40% more likely to be screened than females (aOR 1.37, 95% CI: 1.20–1.57, p <0.001), as were those aged 45 years and above compared to those below 25 years (aOR 1.39, 95% CI: 1.06–1.83, p = 0.017). Individuals with hypertension also had a 33% increased likelihood of screening (aOR 1.33, 95% CI: 1.07–1.65, p = 0.011), and those initiated on non–TDF-based regimens were significantly more likely to be screened (aOR 1.66, 95% CI: 1.09–2.53, p = 0.018). In contrast, having diabetes or HIV-associated opportunistic infections did not affect one’s odds of getting kidney disease screening.

At the urban site, participants with higher CD4 counts (>500 cells/µL) were significantly less likely to be screened compared to those with CD4 <200 cells/µL (aOR 0.69, 95% CI: 0.52– 0.91, p = 0.008, Table 3B). However, having hypertension, diabetes, and opportunistic infections, or initiating a TDF-based ART regimen was not associated with increased odds of kidney disease screening.

## Discussion

In this analysis of data from three large HIV clinics caring for approximately 50,000 PWH in Uganda, we found a low overall prevalence of kidney disease screening at ART initiation. Screening was particularly rare at rural sites where screening rates were less than 10%, and have remained low since 2022, when guidelines officially recommended kidney disease screening for all. Despite the presence of guidelines and low-cost, evidence-based interventions to slow CKD progression, these data would suggest that the health system in Uganda is missing crucial opportunities to improve primary care management for PWH, such as kidney disease screening, identification of CKD and opportunities to improve longterm care of those with the condition.(27)

Our findings support those of other studies, indicating that most patients with kidney disease are not screened and are diagnosed with advanced kidney disease, when treatment options are limited.(28-30) Such low rates of screening are not unique to PWH, but also extend to other high-risk groups in the region. (27, 31, 32) Indeed, only a few countries, such as South Africa and Egypt, have established national screening programs for high-risk populations, and achieved adequate CKD screening rates.(33, 34) The low rates of screening and early detection of reversible kidney disease can be particularly devastating in low- and middle income countries (LMICs), where fragile healthcare systems are already strained, exposing individuals and families to catastrophic treatment costs and poor outcomes associated with advanced kidney disease.(27, 35-39)

There is limited data regarding kidney disease screening for PWH from high-income countries (HICs). A study from Australia reports kidney disease screening rates above 90%.(40) Most kidney disease screening studies from HICs have been among other high-risk populations, such as those with diabetes mellitus and hypertension.(41-43) The reported high rates of screening in these studies highlight the established kidney disease screening programs in HICs. The decline and/or stagnation in CKD screening rates at rural health facilities in recent years, despite updated guidelines and increased advocacy, is particularly concerning. While the coronavirus-2019 (COVID-19) pandemic may have strained health systems there should be visible trends of recovery post the pandemic, which were not observed in this study at rural sites, which serve the majority of PWH in Uganda.

Our analysis of rural Ugandan health facilities reveals that kidney disease screening disproportionately targets patients with traditional risk factors, such as male gender, older age (45+ years), or comorbid hypertension. This suggests that, in resource-limited settings, healthcare providers may be prioritizing screening for individuals perceived to be at highest clinical risk, possibly reflecting older guidelines that targeted screening based on comorbidities rather than current universal recommendations. Intriguingly, individuals initiated on a non-TDF ART regimen were 1.7 times more likely to be screened, perhaps reflecting provider suspicion of underlying renal dysfunction that influenced baseline creatinine testing and regimen selection. Conversely, the large urban health facility in our study demonstrated dramatically improved rates of screening than the rural centers, highlighting how local capacity can shape guideline adherence. At this facility, screening rates increased substantially in the “creatinine for all” era to approximately 80% in the most recent years, reflecting a successful scale-up and implementation of the updated national guidelines. Yet, even here, screening remained selective. Participants with higher CD4 counts (>500 cells/µL) were 30% less likely to be screened than those with advanced HIV (CD4 <200 cells/µL), and those aged ≥45 or with tuberculosis were significantly under-screened. These patterns, especially the lower screening rate among older adults and those with TB suggest clinical triage based on perceived acuity, despite guidelines and known renal risks.(24, 44, 45) Even in well-resourced settings, provider discretion and patient characteristics continue to shape screening decisions more than guidelines alone. These screening trends may also reflect gaps in knowledge and awareness of guideline updates, underscoring the need for emphasis on unique risk factors such as HIV and ART in high-burden settings.^28,(46-50)^ Systemic health sector constraints, such as unreliable laboratory supply chains and stock-outs in rural areas, may also be contributing.

An important area for future research will be to explore the factors that facilitate and restrict kidney disease screening in the rural health facilities of HIV care in the region. This information may help inform strategies to increase routine screening in practice.In addition, clinics could implement quality improvement methods to track and report kidney disease screening as well as implementing electronic health records’ alerts if creatinine is deranged. This information may help clinicians to identify PWH and kidney disease to facilitate early intervention.

Our study has several limitations. Firstly, data were derived from clinical databases and may not reflect or capture all cases of kidney disease screening. For instance, due to the absence of a national health insurance scheme and amidst frequent stock-outs of diagnostic supplies, some participants may have undergone screening tests at private facilities, which may not be captured in routine clinical data systems. Additionally, data clerks manually enter results after they are printed as hard copies, potentially leading to an underestimation of true kidney disease screening rates. Secondly, we were unable to obtain data on key contextual variables that may have contributed to the observed low screening rates, such as the availability of diagnostic resources at each facility or the composition and training levels of healthcare providers involved in ART initiation during the study period. As a result, we cannot report on the influence of these factors on screening practices. Nevertheless, we present data from three large public-sector HIV clinics in Uganda, spanning urban and rural settings and reflecting routine clinical practice and use of programmatic data that includes a broad patient population, enhancing the generalizability of our findings to comparable settings.

In summary, we found a low overall prevalence of kidney disease screening for PWH at ART initiation in Uganda, particularly at rural sites where screening rates remain below 10%. These findings provide insights for health system strengthening for kidney disease in the region, where care delivery and adherence to clinical guidelines are often influenced by factors such as donor support, diagnostic resource availability, and local infrastructure constraints. Future research should investigate the underlying reasons for low and selective kidney disease screening, especially in rural areas. Implementation science approaches may help identify health system, provider, and patient-level barriers influencing screening uptake. Prospective studies using both serum creatinine and urine albumin creatinine ratio are also needed to better quantify CKD burden in HIV populations in Uganda and the region, to ensure the optimization of the health and well-being of this high-risk population.

## Data Availability

All data used in this manuscript preparation is available from the corresponding author upon reasonable request.

## Notes

**Funding:** This Research project was supported by the Fogarty International Center of the National Institutes of Health (NIH) under Award Number D43TW010543. MJS acknowledges additional support from the NIH (K24 HL166024). This article has arisen, in whole or in part by the National Institutes of Health (NIH), and is subject to the NIH’s Plan to Enhance Public Access to the Results of NIH-Supported Research [2024 NIH Public Access Policy], NOT- OD-25-047. As a result, the author retains the rights necessary to comply with this policy, including the right to submit, or have submitted to the National Library of Medicine’s PubMed Central an electronic version of the final, peer-reviewed manuscript to be made available in PubMed Central on the official date of publication. The content is solely the responsibility of the authors and does not necessarily represent the official views of the National Institutes of Health.

**Conflicts of Interest:** All authors report no conflict of interest.

### Competing Interest Statement

The authors have declared no competing interest.

### Funding Statement

This Research project was supported by the Fogarty International Center of the National Institutes of Health (NIH) under Award Number D43TW010543. MJS acknowledges additional support from the NIH (K24 HL166024). This article has arisen, in whole or in part by the National Institutes of Health (NIH), and is subject to the NIH's Plan to Enhance Public Access to the Results of NIH-Supported Research [2024 NIH Public Access Policy], NOT-OD-25-047. As a result, the author retains the rights necessary to comply with this policy, including the right to submit, or have submitted to the National Library of Medicine’s PubMed Central an electronic version of the final, peer-reviewed manuscript to be made available in PubMed Central on the official date of publication. The content is solely the responsibility of the authors and does not necessarily represent the official views of the National Institutes of Health.

